# Rates of Positive M-CHAT-R Screenings by Pandemic Birth and Prenatal SARS-CoV-2 Exposure

**DOI:** 10.1101/2024.02.20.24302892

**Authors:** Morgan R. Firestein, Angela Gigliotti Manessis, Jen Warmingham, Yunzhe Hu, Morgan A. Finkel, Margaret Kyle, Maha Hussain, Imaal Ahmed, Andréane Lavallée, Ana Solis, Vitoria Chaves, Cynthia Rodriguez, Sylvie Goldman, Rebecca A. Muhle, Seonjoo Lee, Judy Austin, Wendy G. Silver, Kally C. O’Reilly, Jennifer M. Bain, Anna A. Penn, Jeremy Veenstra-VanderWeele, Melissa S. Stockwell, William P. Fifer, Rachel Marsh, Catherine Monk, Lauren C. Shuffrey, Dani Dumitriu

**Author notes:** Corresponding Author: Dani Dumitriu, M.D., Ph.D., New York State Psychiatric Institute Columbia University Irving Medical Center 1051 Riverside Drive, Suite 4807, New York, NY 10032. Dr. Firestein and Ms. Gigliotti Manessis contributed equally to this article.

## Abstract

Maternal stress and viral illness during pregnancy are associated with neurodevelopmental conditions in offspring. Children born during the COVID-19 pandemic, including those exposed prenatally to maternal SARS-CoV-2 infections, are reaching the developmental age for the assessment of risk for neurodevelopmental conditions. We examined associations between birth during the COVID-19 pandemic, prenatal exposure to maternal SARS-CoV-2 infection, and rates of positive screenings on the Modified Checklist for Autism in Toddlers-Revised (M-CHAT-R). Data were drawn from the COVID-19 Mother Baby Outcomes (COMBO) Initiative. Participants completed the M-CHAT-R as part of routine clinical care (COMBO-EHR cohort) or for research purposes (COMBO-RSCH cohort). Maternal SARS-CoV-2 status during pregnancy was determined through electronic health records. The COMBO-EHR cohort includes n=1664 children (n=442 historical cohort, n=1222 pandemic cohort; n=997 SARS-CoV-2 unexposed prenatally, n=130 SARS-CoV-2 exposed prenatally) who were born at affiliated hospitals between 2018-2023 and who had a valid M-CHAT-R score in their health record. The COMBO-RSCH cohort consists of n=359 children (n=268 SARS-CoV-2 unexposed prenatally, n=91 SARS-CoV-2 exposed prenatally) born at the same hospitals who enrolled into a prospective cohort study that included administration of the M-CHAT-R at 18-months. Birth during the pandemic was not associated with greater likelihood of a positive M-CHAT-R screen in the COMBO-EHR cohort. Maternal SARS-CoV-2 was associated with lower likelihood of a positive M-CHAT-R screening in adjusted models in the COMBO-EHR cohort (OR=0.40, 95% CI=0.22 - 0.68, *p*=0.001), while analyses in the COMBO-RSCH cohort yielded similar but non-significant results (OR=0.67, 95% CI=0.31-1.37, *p*=0.29).These results suggest that children born during the first 18 months of the COVID-19 pandemic and those exposed prenatally to a maternal SARS-CoV-2 infection are not at greater risk for screening positive on the M-CHAT-R.

## Introduction

Since the onset of the COVID-19 pandemic, we have gained substantial knowledge about the SARS-CoV-2 virus and resulting COVID-19 pathophysiology. However, longitudinal research is needed to determine long-term consequences of early exposure to both the virus and the pandemic environment, especially those affecting the youngest generations.^1–3^ Most reports have provided reassuring results suggesting no association between prenatal exposure to maternal SARS-CoV-2 infection and child neurodevelopment.^4–8^ One research group found that a higher proportion of prenatally exposed male infants received ICD-10 codes for speech disorders at age 12 months, but not at 18 months. This finding, coupled with a lower-than-expected rate of SARS-CoV-2 infections, suggests a high rate of misclassification and false negatives which may limit the interpretability of these results.^9,10^ Further, studies of infants born during previous pandemics have shown that neurodevelopmental needs are frequently unobserved until years later,^11^ necessitating additional follow-up.

Beyond prenatal exposure to SARS-CoV-2, the COVID-19 generation–the generation born during this pandemic–experienced an unusual environment during infancy and early childhood secondary to societal changes and restrictions. Several reports from diverse populations revealed neurodevelopmental differences between pandemic-born compared to pre-pandemic-born children.^4,6,7,12^

The COVID-19 pandemic therefore presents a two-pronged mechanism through which the COVID-19 generation may be at greater risk for neurodevelopmental conditions such as autism spectrum disorders.^6,13^ Maternal prenatal psychological distress has been implicated in increased neurodevelopmental risk through a fetal programming framework.^14,15^ Separately, maternal immune activation (MIA) is among the potential mechanisms through which prenatal exposure to SARS-CoV-2 has been hypothesized to impact offspring neurodevelopment. Previous outbreaks of human coronaviruses (such as SARS and Middle East Respiratory Syndrome) indicate that severe infection during pregnancy may negatively affect both maternal health and infant well-being, possibly through MIA.^13,16^ Based on these premises, we hypothesized that infants exposed to a maternal SARS-CoV-2 in utero would be at increased risk of screening positive on the M-CHAT-R.

Given the benefits of early intervention,^17,18^ special attention should be directed towards evaluating the relationship between fetal exposure to maternal SARS-CoV-2 infection, the COVID-19 pandemic environment, and child neurodevelopment. Children who were *in utero* during peak periods of the pandemic are reaching the age at which early indicators of risk for autism may emerge. We compared scores on the parent-report Modified Checklist for Autism in Toddlers-Revised (M-CHAT-R) screener between children born before and during the pandemic and those with and without prenatal SARS-CoV-2 exposure. Data were collected as part of the COVID-19 Mother Baby Outcomes (COMBO) Initiative and include clinical M-CHAT-R scores recorded in electronic health records (EHR) and M-CHAT-R scores obtained for research purposes from children born at a large university-based medical center in New York City between January 2018 and September 2021.

## Methods

### Study Design and Participants

Data were drawn from the COVID-19 Mother Baby Outcomes (COMBO) Initiative, which includes children born at Columbia University Irving Medical Center (CUIMC)-affiliated NewYork-Presbyterian (NYP) Morgan Stanley Children’s Hospital and NYP Allen Pavilion Hospital in New York City between January 2018 and September 2021. COMBO includes an electronic health record (EHR) arm (COMBO-EHR cohort) with a waiver of consent and a prospective research arm (COMBO-RSCH cohort) with on-going enrollment and consent of mother-infant dyads (eFigure 1). Across the two cohorts, demographic and clinical variables, including maternal SARS-CoV-2 status during pregnancy, were acquired through a combination of automated abstraction and manual review of the participants’ EHRs. As part of routine clinical care, all CUIMC/NYP-affiliated pediatric clinics began universally administering the M-CHAT-R between 16-30 months of age in February 2020, and scores are recorded in the EHR. Separately, mothers enrolled in COMBO-RSCH are asked to complete the M-CHAT-R at the 18-month study follow-up for research purposes. All study procedures have been reviewed and approved by the CUIMC Institutional Review Board. This study followed the Strengthening the Reporting of Observational Studies in Epidemiology (STROBE) reporting guidelines.

**Figure 1.**
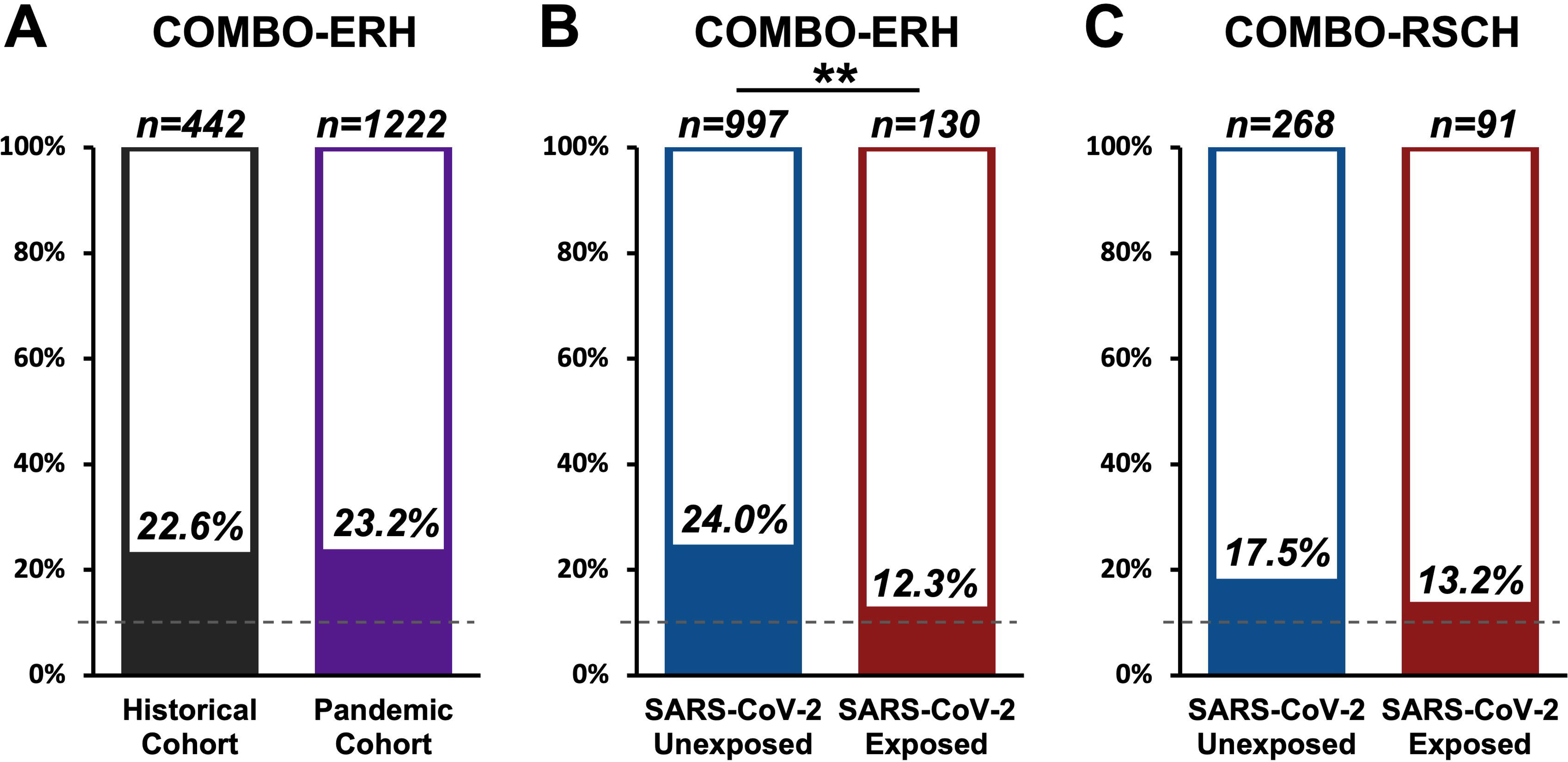
Percent of Children Screening Positive on the M-CHAT-R in the COMBO-EHR and COMBO-RSCH Datasets. Dashed grey lines indicate previously reported rates of positivity in low-risk, US samples.

### COMBO-EHR Cohort

We identified n=1747 children born in the NYP hospital system during the study period with clinical M-CHAT-R scores in the EHRs. For children with more than one M-CHAT-R score, we used the most recent score obtained within the valid age range. We excluded n=80 children because their only M-CHAT-R score was obtained outside the range of 16-30 months. We excluded an additional n=3 children who received a total M-CHAT-R score of 17 that resulted from all items being marked as “yes,” suggesting invalid parental report. In total, we included n=1664 children in COMBO-EHR analyses.

### COMBO-RSCH Cohort

At the time of this analysis, the COMBO Initiative had enrolled n=907 mother-infant dyads who had been admitted to the labor & delivery unit and/or well-baby/NICU units of the CUIMC-NYP hospital system in New York City during the study period. Overall, n=421 enrolled mothers completed the M-CHAT-R for research purposes as part of a REDCap survey administered at the 18-month study time point. Sixty-two children were excluded from COMBO-RSCH analyses because they had M-CHAT-R scores obtained for both clinical and research purposes. These cases were only included in COMBO-EHR dataset to prioritize the analysis of clinical screening data. In total, we included n=359 children in COMBO-RSCH analyses.

### Timing of Birth Relative to the COVID-19 Pandemic

The COMBO-EHR cohort includes n=442 infants born prior to the COVID-19 pandemic (historical cohort) with dates of birth between January 2018–February 2020 and n=1222 infants born during the COVID-19 pandemic (pandemic cohort) with dates of birth between March 2020–September 2021. In contrast, all infants in the COMBO-RSCH cohort were born during the COVID-19 pandemic. Therefore, the historical versus pandemic birth comparison could only be conducted on the COMBO-EHR cohort. Of note, all infants in the COMBO-EHR cohort, including those born prior to the pandemic (historical cohort), were assessed *during* the pandemic as universal M-CHAT-R screening at pediatric clinics coincided with the onset of the pandemic for unrelated reasons. Thus, every child in the COMBO-EHR cohort had *postnatal* exposure to the pandemic environment.

### Determination of Prenatal Maternal SARS-CoV-2 Infection

Maternal SARS-CoV-2 status during pregnancy was determined through EHR abstraction and review. Detailed descriptions of our methods for determining maternal SARS-CoV-2 status during pregnancy have been reported previously^4,5^ and are outlined in eFigure 2. Briefly, on March 22, 2020, CUIMC began universal testing of all delivering patients for SARS-CoV-2 through nasopharyngeal polymerase chain reaction (PCR) and on July 20, 2020, universal serology testing for SARS-CoV-2 antibodies was implemented. Infants born before November 1, 2020, were considered exposed during pregnancy if their mother had a positive PCR and/or serological test during pregnancy or at delivery. If their mother had a negative PCR test and no available serological test result, they were classified as unexposed. For infants born after November 1, 2020, mothers could have had a pre-pregnancy infection that would result in a positive serology test during pregnancy. Therefore, these infants were considered exposed during pregnancy if their mother had a positive PCR and/or antigen test during pregnancy. If their mothers did not have a recorded positive PCR or antigen test but had a positive serological test and COVID-19 symptoms during the pregnancy, they were also considered exposed. Infants were classified as unexposed if their mother had a positive serology test during pregnancy or at the time of delivery, but had no symptoms during the pregnancy, had negative serological testing, or had no serology test results and no symptoms during the pregnancy determined through manual chart review. We have previously estimated that this approach has a 0.67% false-negative rate (eMethods 1).

### Modified Checklist for Autism in Toddlers (M-CHAT-R)

The M-CHAT-R was developed to assess risk for autism based on parent report of their child’s development between 16-30 months of age and consists of twenty ‘yes’/’no’ items that focus on a child’s milestones as well as early autism symptoms that present between the normed age span.^19^ Total scores are calculated and categorized into ranges of “low-risk” (0-2); “medium-risk” (3-7); “high-risk” (8-20). To improve power, we collapsed across the medium-risk and high-risk ranges to define a medium/high-risk range, which we defined as screening positive.

The M-CHAT-R has been validated as having high sensitivity and specificity as a measure for likelihood of autism, with diagnoses confirmed through a variety of measures such as the Autism Diagnostic Observation Schedule and is widely adopted by clinicians.^20,21^ The M-CHAT-R was administered in either English or Spanish depending on the caregiver’s preference.

### Statistical analysis

Statistical analyses were conducted in R version 4.3.1 (R Foundation for Statistical Computing).^22^ M-CHAT-R scores were obtained through EHR review for the COMBO-EHR cohort and using a forced-choice online survey for the COMBO-RSCH cohort, resulting in no missingness of data. Since participants in both cohorts were born in the same hospital system, all demographic variables and other covariates were obtained via the EHRs. For each of our three primary analyses, we performed separate unadjusted Chi-squared tests and multivariable logistic regression models. Using data from the COMBO-EHR cohort, we performed an unadjusted Chi-squared test and an adjusted logistic regression model to assess the association between birth during the COVID-19 pandemic and a positive screening on the M-CHAT-R. For the subset of COMBO-EHR children born during the COVID-19 pandemic and the COMBO-RSCH cohort, we performed an unadjusted Chi-squared test and a multivariable logistic regression model to determine the association between prenatal maternal SARS-CoV-2 status and positive screening on the M-CHAT-R. All multivariable models included as covariates the child’s age at assessment, sex assigned at birth, maternal race and ethnicity, mode of delivery, gestational age at delivery, maternal age at delivery, and insurance type as a proxy for socioeconomic status (Medicaid versus commercial). To examine potential demographic differences between groups, comparisons were performed using Chi-squared tests for binomial variables and Wilcoxon rank sum tests for continuous variables. For the primary analyses, we report Chi-squared results for univariate models and odds ratios (ORs) with 95% confidence intervals (CIs) for multivariable models. For all analyses, we conducted separate subgroup analyses of full-term and preterm children, as well as subgroup analyses of male and female children. For analyses of SARS-CoV-2 exposure within both COMBO-EHR and COMBO-RSCH cohorts, we conducted an additional sensitivity analysis omitting cases without serology testing in the EHR. Finally, we compared the proportion of children screening positive on the M-CHAT-R and the age at assessment across the COMBO-RSCH and COMBO-EHR cohorts using independent samples t-tests and Chi-squared tests. All sensitivity analyses used multivariable models. Significance was set at p<0.05.

## Results

### Cohort Characteristics

The COMBO-EHR cohort included *n=*1664 children, n=442 born before March 2020 (historical cohort) and n=1222 born during the COVID-19 pandemic. In the pandemic-born group, 95 dyads lacked maternal PCR and serology data or had unknown infection timing, resulting in n=997 and n=130 SARS-CoV-2 unexposed and exposed children, respectively.

Characteristics of the mother-child dyads included in the COMBO-EHR cohort are summarized in Table 1. The mean maternal age at delivery was 30 ± 6.24 years. The pandemic cohort differed significantly from the historical cohort in maternal age at delivery (*p*=0.002), self-reported race (χ ^2^=597.91, *df=*6, *p*<0.001), ethnicity ( ^2^=596.41, *df*=3, *p*<0.001), infant gestational age (*p*=0.02), child’s age at M-CHAT-R assessment (*p*<0.01), and infant sex at birth χ =6.97*, df=*1, *p*=0.008) (Table 1). Mothers in the SARS-CoV-2 exposed cohort differed significantly from the unexposed cohort in their self-reported race ( ^2^=8.63, *df=*5, *p*=0.12) and ethnicity ( ^2^=12.31, *df=*2, *p*=0.002) (Table 1). Group differences were accounted for in all fully adjusted models.

**Table 1.**
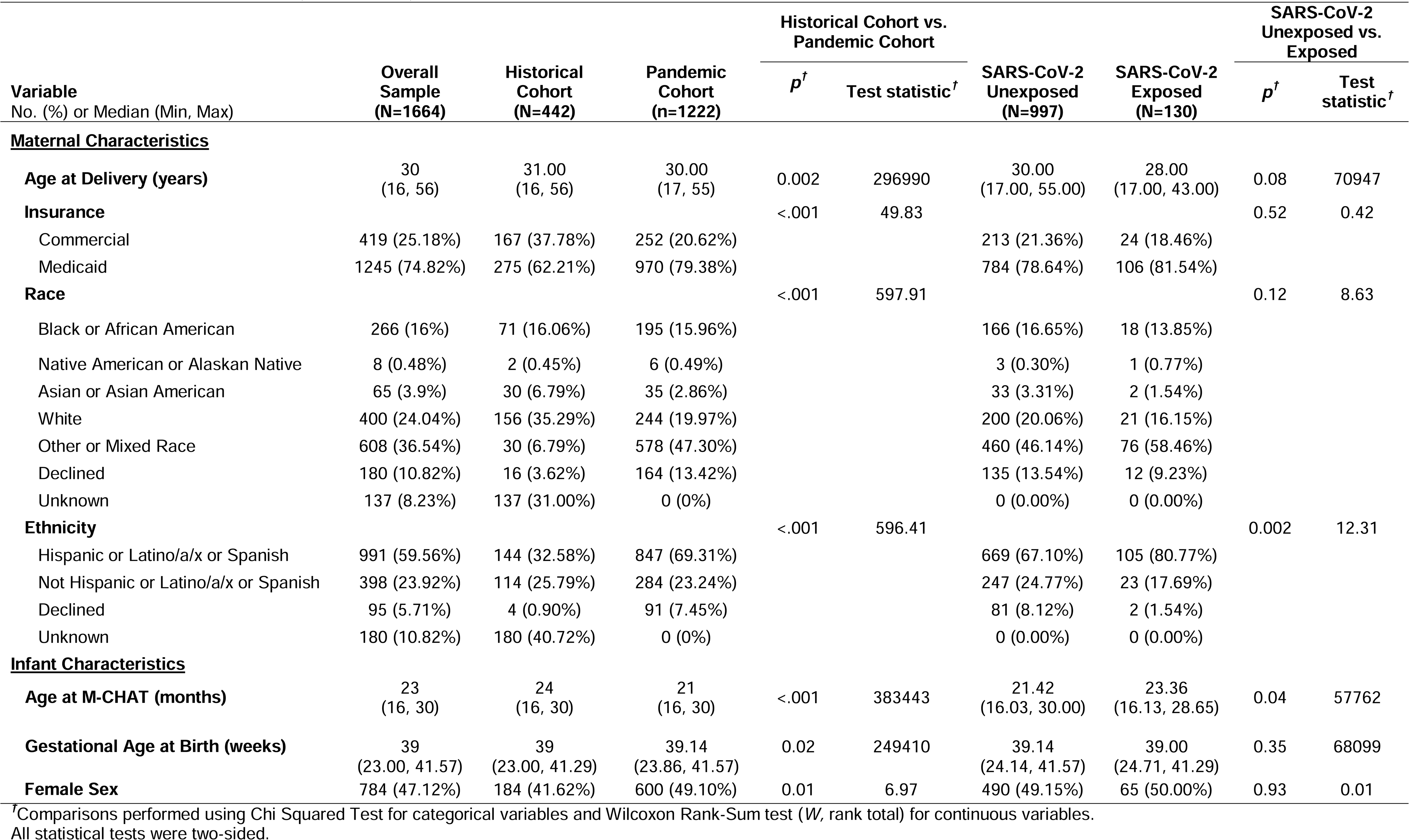
Electronic Medical Record (COMBO-EHR) Cohort Characteristics.

| Variable<br>No. (%) or Median (Min, Max) | Overall<br>Sample<br>(N=1664) | Historical<br>Cohort<br>(N=442) | Pandemic<br>Cohort<br>(n=1222) | Historical Cohort vs.<br>Pandemic Cohort |  | SARS-CoV-2<br>Unexposed<br>(N=997) | SARS-CoV-2<br>Exposed<br>(N=130) | SARS-CoV-2<br>Unexposed vs.<br>Exposed |  |
| --- | --- | --- | --- | --- | --- | --- | --- | --- | --- |
| | | | | $p^{\dagger}$ | Test statistic$^{\dagger}$ | | | $p^{\dagger}$ | Test<br>statistic$^{\dagger}$ |
| **<u>Maternal Characteristics</u>** |  |  |  |  |  |  |  |  |  |
| Age at Delivery (years) | 30<br>(16, 56) | 31.00<br>(16, 56) | 30.00<br>(17, 55) | 0.002 | 296990 | 30.00<br>(17.00, 55.00) | 28.00<br>(17.00, 43.00) | 0.08 | 70947 |
| Insurance |  |  |  | <.001 | 49.83 |  |  | 0.52 | 0.42 |
| Commercial | 419 (25.18%) | 167 (37.78%) | 252 (20.62%) |  |  | 213 (21.36%) | 24 (18.46%) |  |  |
| Medicaid | 1245 (74.82%) | 275 (62.21%) | 970 (79.38%) |  |  | 784 (78.64%) | 106 (81.54%) |  |  |
| Race |  |  |  | <.001 | 597.91 |  |  | 0.12 | 8.63 |
| Black or African American | 266 (16%) | 71 (16.06%) | 195 (15.96%) |  |  | 166 (16.65%) | 18 (13.85%) |  |  |
| Native American or Alaskan Native | 8 (0.48%) | 2 (0.45%) | 6 (0.49%) |  |  | 3 (0.30%) | 1 (0.77%) |  |  |
| Asian or Asian American | 65 (3.9%) | 30 (6.79%) | 35 (2.86%) |  |  | 33 (3.31%) | 2 (1.54%) |  |  |
| White | 400 (24.04%) | 156 (35.29%) | 244 (19.97%) |  |  | 200 (20.06%) | 21 (16.15%) |  |  |
| Other or Mixed Race | 608 (36.54%) | 30 (6.79%) | 578 (47.30%) |  |  | 460 (46.14%) | 76 (58.46%) |  |  |
| Declined | 180 (10.82%) | 16 (3.62%) | 164 (13.42%) |  |  | 135 (13.54%) | 12 (9.23%) |  |  |
| Unknown | 137 (8.23%) | 137 (31.00%) | 0 (0%) |  |  | 0 (0.00%) | 0 (0.00%) |  |  |
| Ethnicity |  |  |  | <.001 | 596.41 |  |  | 0.002 | 12.31 |
| Hispanic or Latino/a/x or Spanish | 991 (59.56%) | 144 (32.58%) | 847 (69.31%) |  |  | 669 (67.10%) | 105 (80.77%) |  |  |
| Not Hispanic or Latino/a/x or Spanish | 398 (23.92%) | 114 (25.79%) | 284 (23.24%) |  |  | 247 (24.77%) | 23 (17.69%) |  |  |
| Declined | 95 (5.71%) | 4 (0.90%) | 91 (7.45%) |  |  | 81 (8.12%) | 2 (1.54%) |  |  |
| Unknown | 180 (10.82%) | 180 (40.72%) | 0 (0%) |  |  | 0 (0.00%) | 0 (0.00%) |  |  |
| **<u>Infant Characteristics</u>** |  |  |  |  |  |  |  |  |  |
| Age at M-CHAT (months) | 23<br>(16, 30) | 24<br>(16, 30) | 21<br>(16, 30) | <.001 | 383443 | 21.42<br>(16.03, 30.00) | 23.36<br>(16.13, 28.65) | 0.04 | 57762 |
| Gestational Age at Birth (weeks) | 39<br>(23.00, 41.57) | 39<br>(23.00, 41.29) | 39.14<br>(23.86, 41.57) | 0.02 | 249410 | 39.14<br>(24.14, 41.57) | 39.00<br>(24.71, 41.29) | 0.35 | 68099 |
| Female Sex | 784 (47.12%) | 184 (41.62%) | 600 (49.10%) | 0.01 | 6.97 | 490 (49.15%) | 65 (50.00%) | 0.93 | 0.01 |

$^{\dagger}$ Comparisons performed using Chi Squared Test for categorical variables and Wilcoxon Rank-Sum test ( $W$ , rank total) for continuous variables. All statistical tests were two-sided.

The COMBO-RSCH cohort (n=359) demographic characteristics are summarized in Table 2. The mean maternal age at delivery was 32 ± 5.08 years. Mother-child dyads with (n=91) and without (n=268) SARS-CoV-2 exposure during pregnancy were compared on the same sociodemographic characteristics described above. Mothers with a history of SARS-CoV-2 infection during pregnancy differed significantly from unexposed mothers in their self-reported ethnicity ( ^2^=9.16, *df=*3, *p*=0.03). Additionally, exposed children were significantly younger at the time of the M-CHAT-R assessment (*p*=0.03). Group differences were accounted for in all fully adjusted models.

**Table 2.**
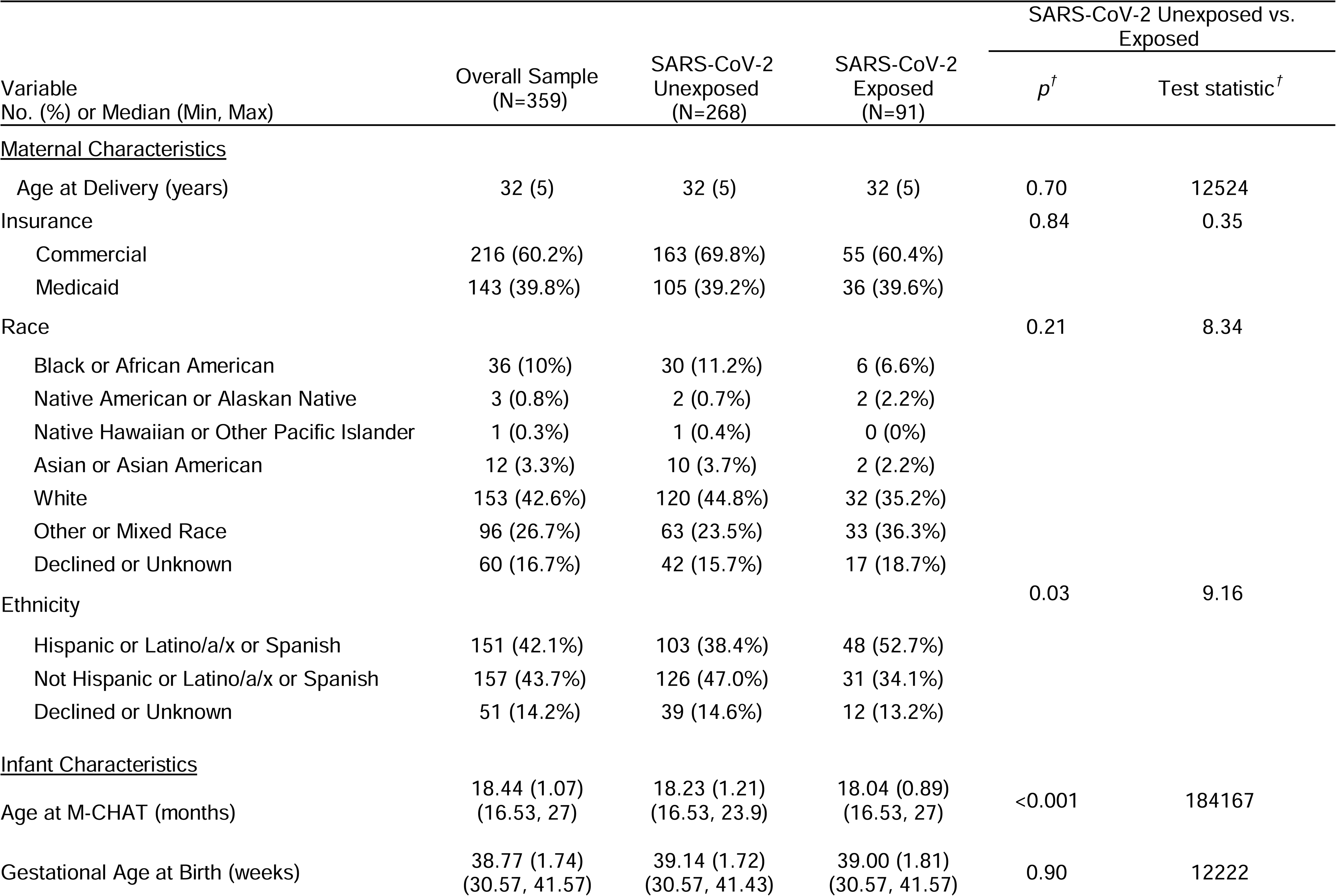
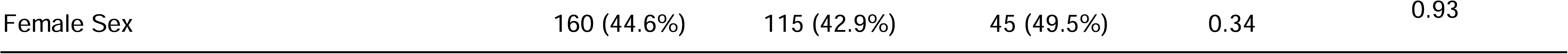
COMBO Research (COMBO-RSCH) Cohort Characteristics.

Additional analyses comparing the COMBO-EHR and COMBO-RSCH cohorts are described in eMethods 2.

### Birth During the Pandemic and M-CHAT-R Positive Screening: COMBO-EHR Cohort

There was no difference in the proportion of children screening positive on the M-CHAT-R between the historical and pandemic cohorts (χ ^2^=0.03, *df=*1, *p=*0.87). Specifically, 22% (n*=*100) of the children in the historical cohort and 23% (n*=*283) of the children in the pandemic cohort screened positive (Figure 1A). The lack of an association between birth during the pandemic and M-CHAT-R positivity was maintained in a fully adjusted model accounting for maternal age, child age at assessment, child sex, race, ethnicity, gestational age, and insurance (*OR*=0.75, *95% CI*=0.52–1.08, *p*=0.12) (Table 3). Similar non-significant findings were observed in adjusted subgroup analyses of preterm (n=204) and full-term subsamples (*n*=1460) (eTable 1) and in adjusted subgroup analyses of male (n=880) and female (n=784) children (eTable 2).

**Table 3.** Comparison of Rates of Positive M-CHAT Screenings Between the Historical (n=442) and Pandemic (n=1222) Cohorts (COMBO-EHR)

| Predictor | B | SE | Odds Ratio | 95% CI | P-value |
| --- | --- | --- | --- | --- | --- |
| Pandemic Birth (*Reference: Born on or after 03/01/2020*) | -0.29 | 0.19 | 0.75 | 0.52 - 1.08 | 0.12 |
| Age at M-CHAT administration (months) | -0.04 | 0.02 | 0.96 | 0.93 - 1.00 | 0.05 |
| Gestational age (weeks) | -0.10 | 0.03 | 0.90 | 0.86 - 0.95 | <.001 |
| Maternal age at delivery >35 | -0.01 | 0.01 | 0.99 | 0.97 - 1.01 | 0.16 |
| Infant sex assigned at birth (Male) | 0.61 | 0.12 | 1.84 | 1.45 - 3.12 | <.001 |
| Insurance (Medicaid) | 0.75 | 0.20 | 2.11 | 1.45 - 3.14 | <.001 |
| Maternal Race–American Indian or Alaska Native | -0.30 | 1.12 | 0.74 | 0.04 - 4.72 | 0.78 |
| Maternal Race–Asian | 0.08 | 0.44 | 1.08 | 0.43 - 2.45 | 0.86 |
| Maternal Race–Black or African American | 0.34 | 0.20 | 1.41 | 0.95 - 2.09 | 0.10 |
| Maternal Race–Declined | 0.29 | 0.23 | 1.33 | 0.83 - 2.10 | 0.22 |
| Maternal Race–Other Combinations Not Described | 0.26 | 0.31 | 1.29 | 0.71 - 2.35 | 0.41 |
| Maternal Race–White | 0.09 | 0.18 | 1.10 | 0.77 - 1.56 | 0.61 |
| Maternal Ethnicity–Not Hispanic or Latino or Spanish Origin | -0.29 | 0.19 | 0.75 | 0.51 - 1.09 | 0.13 |

### Prenatal Maternal SARS-CoV-2 Infection and M-CHAT-R Positive Screenings: COMBO-EHR Cohort

There was a significant association between SARS-CoV-2 exposure during pregnancy and less likelihood of M-CHAT-R positive screenings, which was in the opposite direction as hypothesized (χ ^2^=8.28, *df=*1, *p*=0.004). Specifically, 23% (n=239) of unexposed children and 12% (*n*=16) of exposed children screened positive on the M-CHAT-R (Figure 1B). This association remained significant in a fully adjusted model (OR=0.40, 95% CI=0.22–0.68, *p*=0.004) (Table 4) and persisted in sensitivity analyses excluding cases with elevated potential for misclassification of SARS-CoV-2 exposure due to missing serology testing (n=427, OR=0.43, 95% CI=0.22–0.81, *p*=0.01) (eTable 3). Among full-term children, a lower proportion of children with SARS-CoV-2 exposure screened positive on the M-CHAT-R (n=982, adjusted OR=0.45, 95% CI=0.24–0.79, *p*=0.01), whereas this association did not reach statistical significance among preterm children (n*=*145, OR=0.16, 95% CI=0.01–0.84, *p=*0.07) (eTable 4).

**Table 4.**
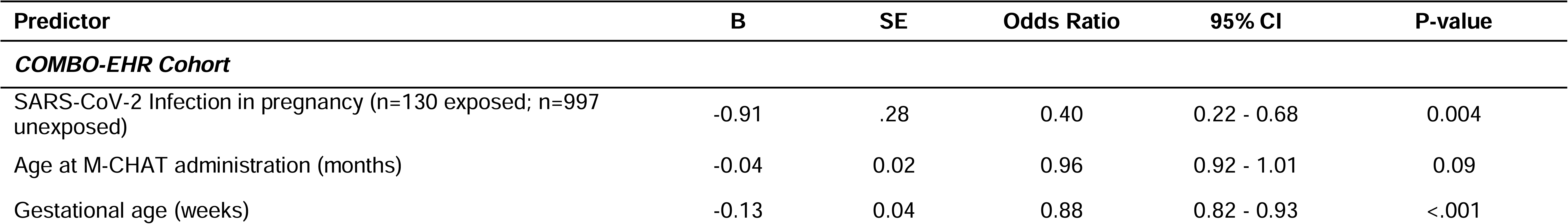
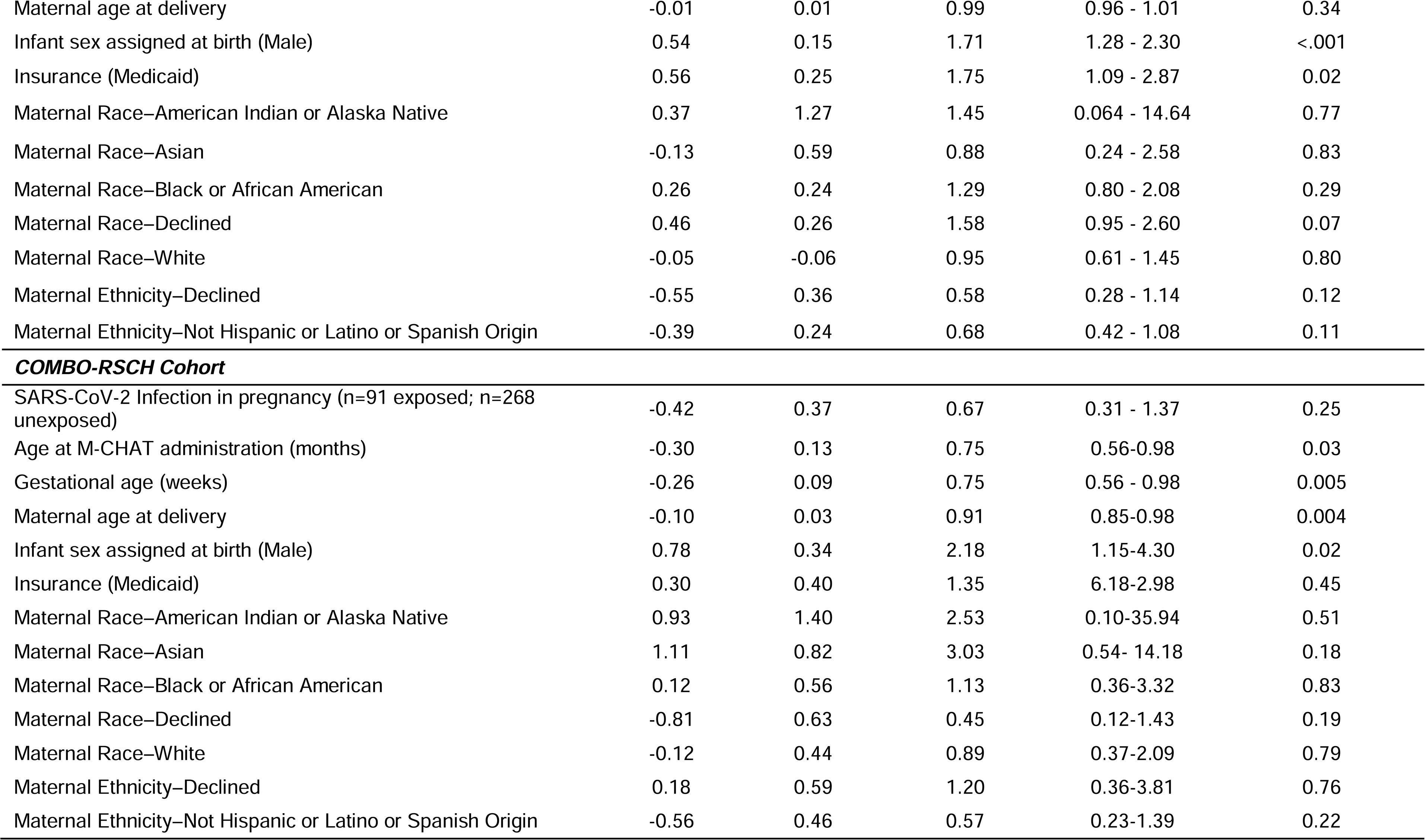
Comparison of Rates of Positive M-CHAT Screenings Between SARS-CoV-2 Exposed and Unexposed Infants Across the COMBO-EHR and COMBO-RSCH Cohorts.

| Predictor | B | SE | Odds Ratio | 95% CI | P-value |
| --- | --- | --- | --- | --- | --- |
| **COMBO-EHR Cohort** |  |  |  |  |  |
| SARS-CoV-2 Infection in pregnancy (n=130 exposed; n=997 unexposed) | -0.91 | .28 | 0.40 | 0.22 - 0.68 | 0.004 |
| Age at M-CHAT administration (months) | -0.04 | 0.02 | 0.96 | 0.92 - 1.01 | 0.09 |
| Gestational age (weeks) | -0.13 | 0.04 | 0.88 | 0.82 - 0.93 | <.001 |
| Maternal age at delivery | -0.01 | 0.01 | 0.99 | 0.96 - 1.01 | 0.34 |
| Infant sex assigned at birth (Male) | 0.54 | 0.15 | 1.71 | 1.28 - 2.30 | <.001 |
| Insurance (Medicaid) | 0.56 | 0.25 | 1.75 | 1.09 - 2.87 | 0.02 |
| Maternal Race–American Indian or Alaska Native | 0.37 | 1.27 | 1.45 | 0.064 - 14.64 | 0.77 |
| Maternal Race–Asian | -0.13 | 0.59 | 0.88 | 0.24 - 2.58 | 0.83 |
| Maternal Race–Black or African American | 0.26 | 0.24 | 1.29 | 0.80 - 2.08 | 0.29 |
| Maternal Race–Declined | 0.46 | 0.26 | 1.58 | 0.95 - 2.60 | 0.07 |
| Maternal Race–White | -0.05 | -0.06 | 0.95 | 0.61 - 1.45 | 0.80 |
| Maternal Ethnicity–Declined | -0.55 | 0.36 | 0.58 | 0.28 - 1.14 | 0.12 |
| Maternal Ethnicity–Not Hispanic or Latino or Spanish Origin | -0.39 | 0.24 | 0.68 | 0.42 - 1.08 | 0.11 |
| **COMBO-RSCH Cohort** |  |  |  |  |  |
| SARS-CoV-2 Infection in pregnancy (n=91 exposed; n=268 unexposed) | -0.42 | 0.37 | 0.67 | 0.31 - 1.37 | 0.25 |
| Age at M-CHAT administration (months) | -0.30 | 0.13 | 0.75 | 0.56-0.98 | 0.03 |
| Gestational age (weeks) | -0.26 | 0.09 | 0.75 | 0.56 - 0.98 | 0.005 |
| Maternal age at delivery | -0.10 | 0.03 | 0.91 | 0.85-0.98 | 0.004 |
| Infant sex assigned at birth (Male) | 0.78 | 0.34 | 2.18 | 1.15-4.30 | 0.02 |
| Insurance (Medicaid) | 0.30 | 0.40 | 1.35 | 6.18-2.98 | 0.45 |
| Maternal Race–American Indian or Alaska Native | 0.93 | 1.40 | 2.53 | 0.10-35.94 | 0.51 |
| Maternal Race–Asian | 1.11 | 0.82 | 3.03 | 0.54- 14.18 | 0.18 |
| Maternal Race–Black or African American | 0.12 | 0.56 | 1.13 | 0.36-3.32 | 0.83 |
| Maternal Race–Declined | -0.81 | 0.63 | 0.45 | 0.12-1.43 | 0.19 |
| Maternal Race–White | -0.12 | 0.44 | 0.89 | 0.37-2.09 | 0.79 |
| Maternal Ethnicity–Declined | 0.18 | 0.59 | 1.20 | 0.36-3.81 | 0.76 |
| Maternal Ethnicity–Not Hispanic or Latino or Spanish Origin | -0.56 | 0.46 | 0.57 | 0.23-1.39 | 0.22 |

Within both female and male subsamples, a lower proportion of children with SARS-CoV-2 exposure screened positive on the M-CHAT-R (females: n=555, adjusted OR=0.30 95% CI=0.10–0.71, *p*=0.01; males: *n=*572, OR=0.48, 95% CI=0.23–0.93, *p*=0.04) (eTable 5).

### Prenatal Maternal SARS-CoV-2 Infection and M-CHAT-R Positive Screenings: COMBO-RSCH Cohort

Overall, fewer SARS-CoV-2 exposed children screened positive (13%, n=12) compared to 17% (n=47) of unexposed children in the COMBO-RSCH cohort (Figure 1C). However, this association was not statistically significant (χ ^2^=0.65, *df=*1, *p*=0.42). The lack of a significant association persisted in a fully adjusted model with the same covariates as those used in the COMBO-EHR analysis (OR=0.67, 95% CI=0.31–1.37, *p*=0.25) (Table 4). Similar non-significant associations were observed in adjusted subgroup analyses of preterm (n=37) and full-term subsamples (n=322) (eTable 6) and in subgroup analyses of male (n*=*199) and female (n*=*160) children (eTable 7).

## Discussion

Considering the conflicting evidence on the relationship between COVID-19 and infant neurodevelopment, continued long-term monitoring of the COVID-19 generation is important for public health and educational policy. We evaluated the neurodevelopmental risk of children born before and during the COVID-19 pandemic and those with and without prenatal SARS-CoV-2 exposure using the M-CHAT-R in a demographically diverse sample in New York City. We found that children born before versus during the pandemic did not have significantly different M-CHAT-R positivity rates. Contrary to our hypothesis, we found that maternal SARS-CoV-2 infection during pregnancy was not associated with increased risk for a positive autism screen based on the M-CHAT-R.

Overall, our sample had higher M-CHAT-R positivity rates compared to other samples (22-23% compared to 9% in a general population).^23^ Participants in our analysis primarily reside in an urban city, where the prevalence of autism diagnoses is higher, potentially due to increased access to healthcare services or environmental exposures unique to urban areas.^24^ Our cohort has a uniquely high percentage of Hispanic participants, a population found to have higher positivity rates on the M-CHAT-R.^25–27^ Further, validity on the Spanish version of the M-CHAT-R was conducted in Spain, which is culturally different from our study sample.^28^

In our sample, maternal SARS-CoV-2 exposure during pregnancy was not associated with increased risk for screening positive on the M-CHAT-R. Research suggests that SARS-CoV-2 may cause MIA, with moderate but temporary changes in cytokine levels in pregnant women^29^ that could be associated with infant neurodevelopmental risk.^10^ Our findings align with other reports of no or limited associations between prenatal SARS-CoV-2 exposure and infant neurodevelopment.^4,5,7,12^ Many of these studies, however, including ours, primarily include participants with mild illness. Additional exploration of the relationship between severity of infection and neurodevelopment is needed.

Of note, the percentage of children who were exposed to SARS-CoV-2 in utero had positivity rates that were closer to the expected rate in a general population; whereas the unexposed children had a higher-than-expected M-CHAT-R positivity rate.^30^ It is possible that unmeasured differences between the exposed and unexposed cohorts in conjunction with the subjective nature of the M-CHAT-R could contribute to the unexpected direction of this association. Data suggest that maternal dispositional variables relate to reporting of their child’s behavior.^31^ Mothers who experienced greater stress and vigilance towards the prevention of SARS-CoV-2 may be less likely to become infected and more likely to monitor and report concerning behaviors through the M-CHAT-R.^32,33^ Additionally, the unexpected positivity rates may be due to limitations of the M-CHAT-R as a research tool. We used only one M-CHAT-R score, without follow-up, for each participant. Finally, it is important to note that all children were assessed during the pandemic and had some amount of postnatal exposure to the pandemic environment. Therefore, our analyses are limited to *prenatal* associations and cannot evaluate associations between *postnatal* experiences and M-CHAT-R positivity.

Our findings suggest that neither SARS-CoV-2 infection nor birth during the pandemic is associated with increased risk for autism. For many, the social and economic impact of the pandemic will persist in the years to come and may continue to impact the developmental trajectories of the COVID-19 generation.^34^ There may also be intergenerational consequences that could worsen existing inequalities.^35^ Furthermore, data suggest that the impact of prenatal exposures may not manifest until higher cognitive functions begin to emerge and mature.^36^ The COVID-19 pandemic has resulted in increased mental health needs amongst pregnant and postpartum individuals,^37^ and higher levels of the stress hormone cortisol during pregnancy is associated with lower offspring educational attainment.^38^ At the same time, the pandemic catalyzed a number of changes in the education system.^39^ Continued monitoring of this generation is important to develop targeted and appropriate policies in education and welfare.

## Data Availability

Data included in this study may be available upon request to the authors.

## Acknowledgements

The authors are grateful for the institutional support provided by the Maternal-Child Research Oversight (MaCRO) Committee and the Departments of Pediatrics, Psychiatry, and Obstetrics and Gynecology at Columbia University Irving Medical Center, which made possible the collection of time-sensitive information at the height of the COVID-19 pandemic prior to the availability of external funding opportunities. The authors wish to thank the entire COVID-19 Mother Baby Outcomes (COMBO) Initiative Team for their collaborative and persevering contributions throughout the COVID-19 pandemic and the Columbia University Irving Medical Center Clinical Data Warehouse. Finally, the authors extend particular gratitude toward the families enrolled in COMBO, whose participation continues to inform our understanding of this unprecedented global event. Dr. Dumitriu has pending reimbursement for consulting for Medela Inc. No funding agency played any role in any of the following: design and conduct of the study; collection, management, analysis, and interpretation of the data; preparation, review, or approval of the manuscript; and decision to submit the manuscript for publication. Dr. Dumitriu had full access to all the data in the study and takes responsibility for the integrity of the data and the accuracy of the data analysis. Drs. Firestein, Warmingham, Shuffrey, Dumitriu, and Ms. Manessis are responsible for the data analysis.

## Notes

### Competing Interest Statement

The authors have declared no competing interest.

### Funding Statement

This study was funded by grant R01MH126531 from the National Institute of Mental Health (Drs Marsh, Monk, and Dumitriu), grant T32MH016434 from the National Institute of Mental Health (Dr Firestein), grant K99HD108389 from the Eunice Kennedy Shriver National Institute of Child Health and Human Development (Dr Firestein), and grant P2CHD058486 from the Eunice Kennedy Shriver National Institute of Child Health and Human Development awarded to the Columbia Population Research Center at Columbia University (Dr Fifer).

### Author Declarations

All procedures were approved by the Columbia University Institutional Review Board or the New York State Psychiatric Institute Institutional Review Board. All necessary patient/participant consent has been obtained and the appropriate institutional forms have been archived.

