## SUPPLEMENTAL APPENDIX for "Rates of Positive M-CHAT-R Screenings by Pandemic Birth and Prenatal SARS-CoV-2 Exposure"

**SUPPLEMENTAL MATERIALS**

**eMethods 1. Estimated misclassification of maternal SARS-CoV-2 status during pregnancy**

The COMBO Initiative utilizes NYP’s Clinical Data Warehouse to automatically extract data, including maternal SARS-CoV-2 test results by PCR and serology, on all delivering mothers and their infants every two weeks. After receiving the data through automated EHR extraction, a detailed chart review was conducted for each SARS-CoV-2 exposed mother/infant dyad to determine symptom severity and timing relative to the pregnancy (e.g., pre-pregnancy, first, second, or third trimester, after delivery). This chart review was primarily conducted by pediatricians and obstetricians providing patient care during the pandemic, or research assistants trained by these clinicians. For women identified as exposed based on serology only, the chart was reviewed for outside records scanned into our EHR system, in person and telehealth visits, and all other notes between onset of the pandemic (early March 2020) and birth. False positive SARS-CoV-2 results by both PCR and serology are extremely rare and most of our patients had either multiple positive tests or experienced symptoms. Therefore, the potential for misclassifying unexposed mothers into the exposed group is very low and was not considered in our exploration of effects of misclassification. For mothers classified as being unexposed to SARS-CoV-2 during pregnancy, there were two sources of potential misclassification: asymptomatic disease and nonseroconversion. The asymptomatic disease in our population has been estimated to be approximately 30%. The most conservative estimate of misclassification of exposed dyads into the unexposed group prior to 7/20/2020 (when universal testing by serology was initiated) is therefore 30% of the 15% of women expected to have COVID-19 disease during pregnancy, which results in 4.5%. After 7/20/2020, misclassification would occur most likely due to asymptomatic disease in nonseroconverters. Therefore, the most conservative rate of misclassification would be 15% of the 4.5% of asymptomatic women or 0.67% of all women initially screened into the unexposed group.

**eMethods 2. Comparing the COMBO-RSCH Cohort and the COMBO-EHR Cohort**

We evaluated potential differences between children in the COMBO-EHR cohort and children in the COMBO-RSCH cohort to further understand the observed discordant results. Independent samples T-tests revealed that children in the COMBO-EHR cohort were significantly older at the time of the MCHAT-R assessment compared to children in the COMBO-RSCH cohort (*t* = 30.24, *p* < 0.01). Overall, children in the COMBO-RSCH cohort had lower MCHAT-R scores than children in the COMBO-EHR cohort (*t* = 5.5245, *p* < 0.01) and Mann Whitney U test confirmed significantly lower proportion of children in the COMBO-RSCH cohort screened positive on the MCHAT-R compared to children in the COMBO-EHR cohort (*W* = 276830, *p* < 0.01).

**eFigure 1. Flowchart of COMBO-EHR and COMBO-RSCH Cohorts**

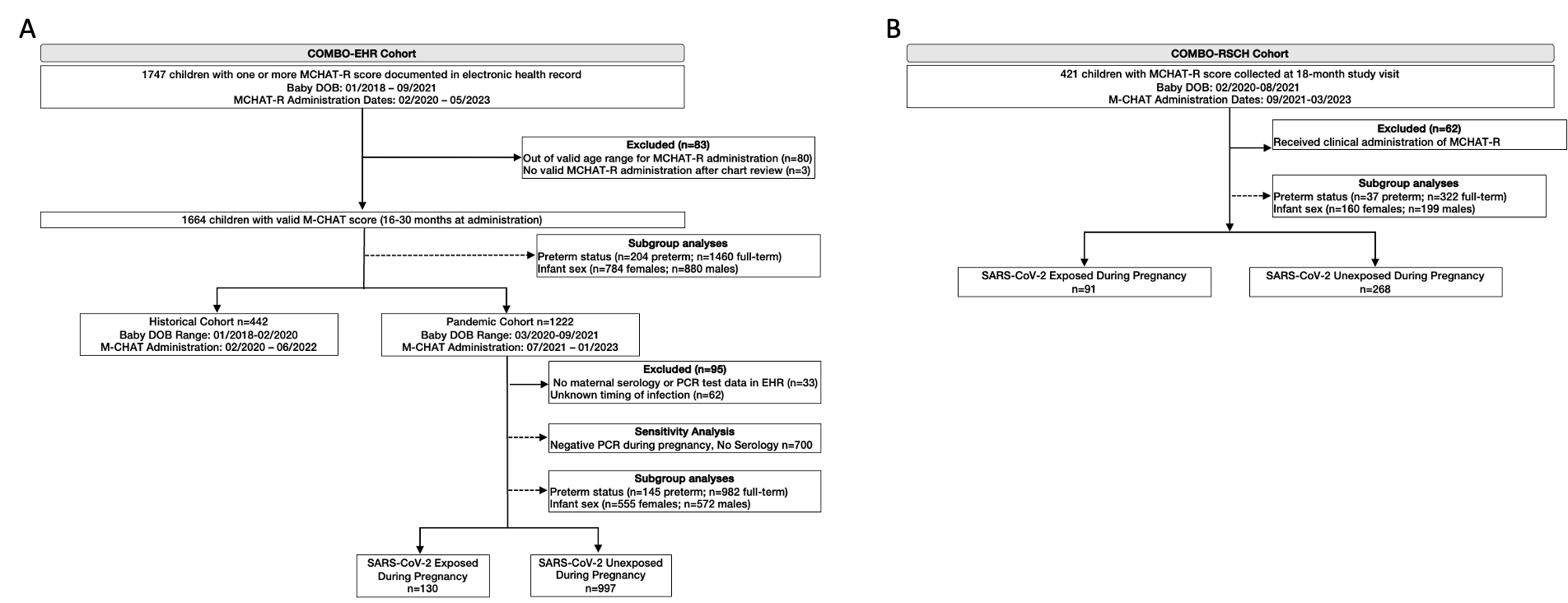

**eFigure 2. SARS-CoV-2 status determination**
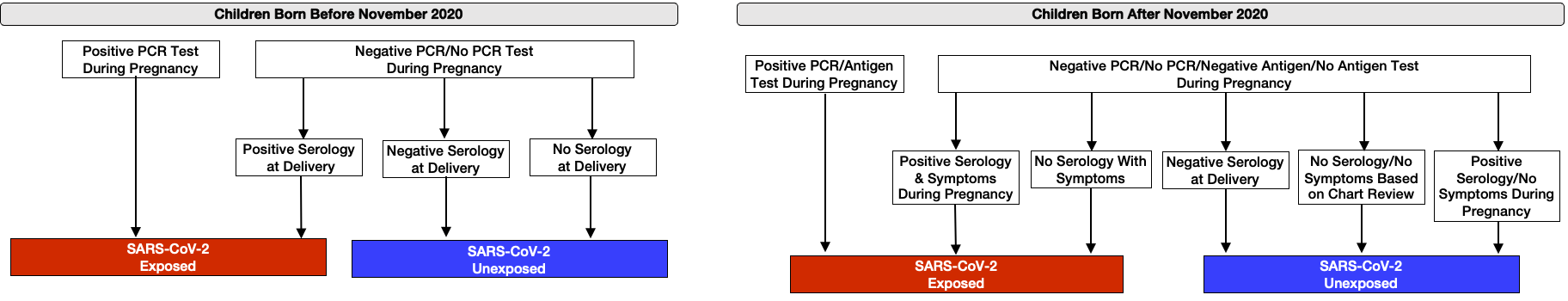

| **eTable 1. Birth Timing and MCHAT-R Positive Screenings in Preterm and Full-Term Subgroups (COMBO-EHR)** | | | | | |
| --- | --- | --- | --- | --- | --- |
| **Predictor** | **B** | **SE** | **Odds Ratio** | **95% CI** | **P-value** |
| ***Preterm Subgroup (n=204)*** |  |  |  |  |  |
| Pandemic Birth (*Reference: Born on or after 03/01/2020*) | 0.06 | .58 | 1.06 | 0.35 - 3.42 | 0.92 |
| Age at M-CHAT administration (months) | 0.04 | 0.051 | 1.04 | 0.94 - 1.15 | 0.43 |
| Maternal age at delivery | -.04 | 0.051 | 0.96 | 0.91 - 1.02 | 0.18 |
| Infant sex assigned at birth (Male) | 0.52 | 0.36 | 1.68 | 0.84 -3.44 | 0.15 |
| Insurance (Medicaid) | 0.45 | 0.54 | 1.58 | 0.56 - 4.78 | 0.40 |
| Maternal Race – American Indian or Alaska Native | 1.10 | 1.49 | 2.99 | 0.11 -84.50 | 0.46 |
| Maternal Race – Asian | -0.68 | 1.23 | 0.51 | 0.02 - 4.24 | 0.58 |
| Maternal Race – Black or African American | 0.36 | 0.57 | 1.44 | 0.46 -4.36 | 0.52 |
| Maternal Race – Declined | -1.03 | 0.77 | 0.35 | 0.06 - 1.42 | 0.18 |
| Maternal Race – Unknown | -0.33 | 1.14 | 0.72 | 0.06 - 6.53 | 0.77 |
| Maternal Race – White | -0.02 | 0.48 | 0.98 | 0.38 - 2.48 | 0.97 |
| Maternal Ethnicity – Declined | 0.25 | 0.99 | 1.29 | 0.14 – 8.65 | 0.80 |
| Maternal Ethnicity – Not Hispanic or Latino | 0.06 | .54 | 1.06 | 0.37 - 3.06 | 0.91 |
| Maternal Ethnicity - Unknown | -1.01 | 1.08 | 0.37 | 0.03 – 2.82 | .35 |
| ***Full-Term Subgroup (n=1460)*** |  |  |  |  |  |
| Pandemic Birth (*Reference: Born on or after 03/01/2020*) | -0.32 | 0.20 | 0.72 | 0.49 - 1.07 | 0.11 |
| Age at M-CHAT administration (months) | -0.05 | 0.02 | 0.95 | 0.92 - 0.99 | 0.02 |
| Maternal age at delivery | -0.01 | 0.01 | 0.99 | 0.97 - 1.02 | 0.54 |
| Infant sex assigned at birth (Male) | 0.64 | 0.13 | 1.89 | 1.46 - 2.46 | <.001 |
| Insurance (Medicaid) | 0.79 | 0.21 | 2.21 | 1.36 - 3.40 | <.001 |
| Maternal Race – American Indian or Alaska Native | -12.60 | 346.59 | 0 | ^a^NA | 0.97 |
| Maternal Race – Asian | 0.19 | 0.47 | 1.21 | 0.45 - 2.93 | 0.69 |
| Maternal Race – Black or African American | 0.34 | 0.22 | 1.41 | 0.92 - 2.15 | 0.11 |
| Maternal Race – Declined | 0.44 | 0.25 | 1.56 | 0.95 - 2.52 | 0.08 |
| Maternal Race – Unknown | 0.34 | 0.32 | 1.41 | 0.75 - 2.63 | 0.28 |
| Maternal Race – White | 0.13 | 0.20 | 1.14 | 0.77 - 1.67 | 0.50 |
| Maternal Ethnicity - Declined | -0.40 | 0.35 | 0.67 | 0.33 – 1.31 | 0.26 |
| Maternal Ethnicity – Not Hispanic or Latino | -0.37 | 0.21 | 0.69 | 0.45 – 1.04 | 0.08 |
| Maternal Ethnicity – Unknown | -0.57 | 0.29 | 0.56 | 0.32 – 0.99 | 0.05 |
| ^a^Estimate of 95% CI lower limit not able to be estimated due to small sample size. | | | | | |

| **eTable 2. Birth Timing and MCHAT-R Positive Screenings in Female and Male Subgroups (COMBO-EHR)** | | | | | |
| --- | --- | --- | --- | --- | --- |
| **Predictor** | **B** | **SE** | **Odds Ratio** | **95% CI** | **P-value** |
| ***Females (n=784)*** |  |  |  |  |  |
| Pandemic Birth (*Reference: Born on or after 03/01/2020*) | -0.35 | 0.31 | 0.71 | 0.39 - 1.32 | 0.26 |
| Age at M-CHAT administration (months) | -0.047 | 0.03 | 0.95 | 0.90 - 1.01 | 0.12 |
| Maternal age at delivery | 0.0087 | 0.017 | 1.01 | 0.98 – 1.04 | 0.61 |
| Insurance (Medicaid) | 0.97 | 0.35 | 2.63 | 1.37 – 5.35 | 0.01 |
| Gestational Age | -0.088 | 0.045 | 0.92 | 0.84 – 1.00 | 0.05 |
| Maternal Race – American Indian or Alaska Native | -12.48 | 487.25 | 0 | ^a^NA | 0.97 |
| Maternal Race – Asian | 0.17 | 0.70 | 1.18 | 0.25 – 4.22 | 0.81 |
| Maternal Race – Black or African American | 0.54 | 0.31 | 1.71 | 0.93 – 3.12 | 0.08 |
| Maternal Race – Declined | 0.24 | 0.36 | 1.27 | 0.61 – 2.50 | 0.51 |
| Maternal Race – Unknown | 0.42 | 0.55 | 1.53 | 0.50 – 4.56 | 0.44 |
| Maternal Race – White | -0.09 | 0.28 | 0.91 | 0.52 – 1.58 | 0.75 |
| Maternal Ethnicity - Declined | -0.09 | 0.50 | 0.91 | 0.33 – 2.32 | 0.85 |
| Maternal Ethnicity – Not Hispanic or Latino | -0.46 | 0.32 | 0.63 | 0.34 – 1.17 | 0.15 |
| Maternal Ethnicity – Unknown | -1.20 | 0.56 | 0.30 | 0.094 – 0.86 | 0.03 |
| ***Males (n=880)*** |  |  |  |  |  |
| Pandemic Birth (*Reference: Born on or after 03/01/2020*) | -0.29 | 0.24 | 0.75 | 0.47 - 1.21 | 0.23 |
| Age at M-CHAT administration (months) | -0.03 | 0.02 | 0.97 | 0.92 – 1.01 | 0.17 |
| Maternal age at delivery | -0.03 | 0.01 | 0.97 | 0.95 – 1.0 | 0.04 |
| Insurance (Medicaid) | 0.65 | 0.25 | 1.91 | 1.20 – 3.13 | 0.008 |
| Gestational Age | -0.11 | 0.03 | 0.90 | 0.84 –0.96 | 0.001 |
| Maternal Race – American Indian or Alaska Native | 0.11 | 1.18 | 1.12 | 0.05 – 8.78 | 0.93 |
| Maternal Race – Asian | 0.06 | 0.57 | 1.07 | 0.32 – 3.06 | 0.91 |
| Maternal Race – Black or African American | 0.19 | 0.27 | 1.21 | 0.71 – 2.04 | 0.48 |
| Maternal Race – Declined | 0.32 | 0.31 | 1.37 | 0.73 – 2.52 | 0.31 |
| Maternal Race – Unknown | 0.20 | 0.37 | 1.22 | 0.58 – 2.55 | 0.59 |
| Maternal Race – White | 0.20 | 0.24 | 1.22 | 0.76 – 1.95 | 0.41 |
| Maternal Ethnicity - Declined | -0.48 | 0.43 | 0.62 | 0.26 – 1.43 | 0.27 |
| Maternal Ethnicity – Not Hispanic or Latino | -0.19 | 0.24 | 0.83 | 0.51 – 1.34 | 0.45 |
| Maternal Ethnicity – Unknown | -0.50 | 0.33 | 0.60 | 0.31 – 1.16 | 0.13 |
| ^a^Estimate of 95% CI lower limit not able to be estimated due to small sample size. | | | | | |

| **eTable 3. Sensitivity Analyses of MCHAT-R Positive Screenings Excluding Cases Without Serology Testing (n=427) (*COMBO-EHR)*** | | | | | |
| --- | --- | --- | --- | --- | --- |
| **Predictor** | **B** | **SE** | **Odds Ratio** | **95% CI** | **P-value** |
| SARS-CoV-2 Infection in Pregnancy | -0.84 | 0.33 | 0.43 | 0.22 – 0.81 | 0.01 |
| Age at M-CHAT administration (months) | -0.07 | 0.04 | 0.94 | 0.86 – 1.02 | 0.13 |
| Gestational age (weeks) | -0.19 | 0.05 | 0.83 | 0.75 – 0.92 | <.001 |
| Maternal age at delivery | -0.04 | 0.02 | 0.96 | 0.92 – 1.00 | 0.06 |
| Infant sex assigned at birth (Male) | 0.76 | 0.28 | 2.13 | 1.25 – 3.71 | 0.006 |
| Insurance (Medicaid) | -0.21 | 0.36 | 0.81 | 0.40 – 1.66 | 0.56 |
| Maternal Race – American Indian or Alaska Native | -11.95 | 607.54 | 0 | ^a^NA | 0.98 |
| Maternal Race – Asian | -0.004 | 0.89 | 1.00 | 0.13 – 4.84 | 0.99 |
| Maternal Race – Black or African American | 0.90 | 0.41 | 2.45 | 1.08 – 5.52 | 0.03 |
| Maternal Race – Declined | 0.67 | 0.48 | 1.95 | 0.74 – 4.92 | 0.16 |
| Maternal Race – White | -0.18 | 0.40 | 0.83 | 0.37 – 1.79 | 0.65 |
| Maternal Ethnicity – Declined | -1.03 | 0.65 | 0.36 | 0.09 – 1.22 | 0.11 |
| Maternal Ethnicity – Not Hispanic or Latino or Spanish Origin | -1.05 | 0.40 | 0.35 | 0.16 - 0.76 | 0.009 |
| ^a^Estimate of 95% CI lower limit not able to be estimated due to small sample size. | | | | | |

| **eTable 4. SARS-CoV-2 Exposure and MCHAT-R Positive Screenings in Preterm and Full-Term Subgroup (*COMBO-EHR)*** | | | | | |
| --- | --- | --- | --- | --- | --- |
| **Predictor** | **B** | **SE** | **Odds Ratio** | **95% CI** | **P-value** |
| ***Preterm Subgroup (n=145)*** |  |  |  |  |  |
| SARS-CoV-2 Infection in Pregnancy | -1.83 | 1.00 | 0.16 | 0.01 - 0.84 | 0.07 |
| Age at M-CHAT administration (months) | 0.11 | 0.07 | 1.11 | 0.97 - 1.28 | 0.12 |
| Gestational age (weeks) | -0.43 | 0.12 | 0.65 | 0.50 - 0.81 | <.001 |
| Maternal age at delivery | -0.05 | 0.04 | 0.95 | 0.88 - 1.02 | 0.16 |
| Infant sex assigned at birth (Male) | 0.19 | 0.46 | 1.20 | 0.49 - 3.02 | 0.69 |
| Insurance (Medicaid) | -0.18 | 0.74 | 0.84 | 0.20 - 3.75 | 0.81 |
| Maternal Race – American Indian or Alaska Native | 1.44 | 1.52 | 4.23 | 0.14 - 123.92 | 0.34 |
| Maternal Race – Asian | -1.40 | 1.39 | 0.25 | 0.01 - 3.06 | 0.32 |
| Maternal Race – Black or African American | 0.57 | 0.76 | 1.77 | 0.39 - 8.01 | 0.45 |
| Maternal Race – Declined | -1.29 | 0.92 | 0.28 | 0.03 - 1.40 | .16 |
| Maternal Race – White | -0.66 | 0.66 | 0.52 | 0.13 - 1.80 | 0.32 |
| Maternal Ethnicity – Declined | 0.30 | 1.13 | 1.35 | 0.13 - 12.54 | 0.79 |
| Maternal Ethnicity – Not Hispanic or Latino or Spanish Origin | -0.24 | 0.75 | 0.78 | 0.17 - 3.42 | 0.75 |
| ***Full-Term Subgroup (n=982)*** |  |  |  |  |  |
| SARS-CoV-2 Infection in Pregnancy | -0.80 | 0.30 | 0.45 | 0.24 – 0.79 | 0.01 |
| Age at M-CHAT administration (months) | -0.05 | 0.02 | 0.95 | 0.90 – 0.99 | 0.03 |
| Gestational age (weeks) | -0.12 | 0.08 | 0.88 | 0.76 – 1.02 | 0.10 |
| Maternal age at delivery | -0.004 | 0.01 | 1.00 | 0.97 – 1.02 | 0.77 |
| Infant sex assigned at birth (Male) | 0.59 | 0.16 | 1.80 | 1.31 – 2.47 | <.001 |
| Insurance (Medicaid) | 0.58 | 0.27 | 1.78 | 1.06 – 3.08 | 0.03 |
| Maternal Race – American Indian or Alaska Native | -10.92 | 377.83 | 0 | ^a^NA | 0.98 |
| Maternal Race – Asian | -0.04 | 0.68 | 0.96 | 0.21 – 3.25 | 0.96 |
| Maternal Race – Black or African American | 0.23 | 0.27 | 1.26 | 0.74 – 2.11 | 0.39 |
| Maternal Race – Declined | 0.70 | 0.28 | 2.01 | 1.17 – 3.44 | 0.01 |
| Maternal Race – White | -0.01 | 0.24 | 0.99 | 0.62 – 1.56 | 0.96 |
| Maternal Ethnicity – Declined | -0.77 | 0.39 | 0.46 | 0.21 – 0.97 | 0.05 |
| Maternal Ethnicity – Not Hispanic or Latino or Spanish Origin | -0.49 | 0.27 | 0.61 | 0.36 – 1.02 | 0.06 |
| ^a^Estimate of 95% CI lower limit not able to be estimated due to small sample size. | | | | | |

| **eTable 5. SARS-CoV-2 Exposure and MCHAT-R Positive Screenings in Female and Male Subgroups (*COMBO-EHR)*** | | | | | |
| --- | --- | --- | --- | --- | --- |
| **Predictor** | **B** | **SE** | **Odds Ratio** | **95% CI** | **P-value** |
| ***Females (n=555)*** |  |  |  |  |  |
| SARS-CoV-2 Infection in Pregnancy | -1.23 | 0.49 | 0.30 | 0.10 - 0.71 | 0.01 |
| Age at M-CHAT administration (months) | -0.06 | 0.04 | 0.95 | 0.88 - 1.01 | 0.12 |
| Gestational age (weeks) | -0.13 | 0.06 | 0.88 | 0.79 - 0.99 | 0.02 |
| Maternal age at delivery | 0.01 | 0.02 | 1.01 | 0.97 - 1.05 | 0.56 |
| Insurance (Medicaid) | 0.79 | 0.41 | 2.20 | 1.01 - 5.19 | 0.06 |
| Maternal Race – American Indian or Alaska Native | -12.96 | 596.80 | 0 | ^a^NA | 0.98 |
| Maternal Race – Asian | -0.60 | 1.11 | 0.55 | 0.03 – 3.37 | 0.59 |
| Maternal Race – Black or African American | 0.35 | 0.36 | 1.42 | 0.68 – 2.87 | 0.33 |
| Maternal Race – Declined | 0.53 | 0.39 | 1.69 | 0.76 – 3.60 | 0.18 |
| Maternal Race – White | -0.10 | 0.33 | 0.91 | 0.46 – 1.71 | 0.77 |
| Maternal Ethnicity – Declined | -0.43 | 0.56 | 0.65 | 0.20 – 1.86 | 0.44 |
| Maternal Ethnicity – Not Hispanic or Latino or Spanish Origin | -0.48 | 0.39 | 0.62 | 0.28 – 1.31 | 0.21 |
| ***Males (n=572)*** |  |  |  |  |  |
| SARS-CoV-2 Infection in Pregnancy | -0.73 | 0.36 | 0.48 | 0.23 - 0.93 | 0.04 |
| Age at M-CHAT administration (months) | -0.03 | 0.03 | 0.97 | 0.92 - 1.03 | 0.31 |
| Gestational age (weeks) | -0.14 | 0.05 | 0.87 | 0.80 - 0.95 | 0.002 |
| Maternal age at delivery | -0.03 | 0.02 | 0.97 | 0.94 - 1.00 | 0.10 |
| Insurance (Medicaid) | 0.48 | 0.31 | 1.62 | 0.89 - 3.05 | 0.13 |
| Maternal Race – American Indian or Alaska Native | 1.57 | 1.65 | 4.82 | 0.14 – 168.94 | 0.34 |
| Maternal Race – Asian | 0.14 | 0.73 | 1.15 | 0.23 – 4.39 | 0.85 |
| Maternal Race – Black or African American | 0.14 | 0.33 | 1.15 | 0.59 – 2.18 | 0.68 |
| Maternal Race – Declined | 0.40 | 0.34 | 1.50 | 0.76 – 2.91 | 0.23 |
| Maternal Race – White | -0.02 | 0.29 | 0.98 | 0.54 – 1.74 | 0.95 |
| Maternal Ethnicity – Declined | -0.59 | 0.47 | 0.56 | 0.22 – 1.36 | 0.21 |
| Maternal Ethnicity – Not Hispanic or Latino or Spanish Origin | -0.28 | 0.31 | 0.75 | 0.41 – 1.37 | 0.36 |
| ^a^Estimate of 95% CI lower limit not able to be estimated due to small sample size. | | | | | |

| **eTable 6. SARS-CoV-2 Exposure and MCHAT-R Positive Screenings in Preterm and Full-Term Subgroup (*COMBO-RSCH)*** | | | | | |
| --- | --- | --- | --- | --- | --- |
| **Predictor** | **B** | **SE** | **Odds Ratio** | **95% CI** | **P-value** |
| ***Preterm Subgroup (n=37)*** |  |  |  |  |  |
| SARS-CoV-2 Infection in pregnancy | -1.20 | 1.48 | 0.30 | 0.01- 4.91 | 0.42 |
| Age at M-CHAT administration (months) | 0.35 | 0.71 | 1.41 | 0.34- 6.55 | 0.62 |
| Maternal age at delivery | 0.31 | 0.23 | 1.36 | 0.92-2.38 | 0.17 |
| Infant sex assigned at birth (Male) | 0.40 | 1.60 | 1.49 | 0.07-52.4 | 0.80 |
| Insurance (Medicaid) | 4.51 | 2.38 | 91.00 | 2.16- 41028.3 | 0.06 |
| Maternal Race – American Indian or Alaska Native | 16.90 | 3956.18 | 0 | ^a^NA | 0.10 |
| Maternal Race – Black or African American | -0.60 | 3.10 | 0.57 | 0  -319.52 | 0.90 |
| Maternal Race – White | 0.83 | 2.96 | 2.30 | 0.01-1169.8 | 0.78 |
| Maternal Ethnicity – Not Hispanic or Latino | 0.22 | 1.82 | 1.24 | 0.03-70.4 | 0.90 |
| ***Full-Term Subgroup (n=322)*** |  |  |  |  |  |
| SARS-CoV-2 Infection in pregnancy | -0.32 | 0.39 | 0.72 | 0.32-1.53 | 0.41 |
| Age at M-CHAT administration (months) | -0.34 | 0.15 | 0.71 | 0.51-0.94 | 0.02 |
| Maternal age at delivery | -0.11 | 0.04 | 0.90 | 0.83-0.96 | 0.003 |
| Infant sex assigned at birth (Male) | 0.59 | 0.35 | 1.79 | 0.92-3.64 | 0.09 |
| Insurance (Medicaid) | -0.12 | 0.43 | 0.90 | 0.38-2.07 | 0.78 |
| Maternal Race – American Indian or Alaska Native | -13.2 | 1023.67 | 0 | ^a^NA | 0.99 |
| Maternal Race – Asian | 0.93 | 0.83 | 2.53 | 0.44-1.22 | 0.26 |
| Maternal Race – Black or African American | 0.17 | 0.61 | 1.18 | 0.34-3.84 | 0.78 |
| Maternal Race – Declined | -0.83 | 0.61 | 0.44 | 0.12-1.39 | 0.18 |
| Maternal Race – White | -0.18 | 0.47 | 0.83 | 0.33-2.06 | 0.69 |
| Maternal Ethnicity - Declined | 0.04 | 0.59 | 1.04 | 0.31-3.32 | 0.95 |
| Maternal Ethnicity – Not Hispanic or Latino | -0.68 | 0.50 | 0.51 | 0.18-1.34 | 0.17 |
| Maternal Ethnicity – Unknown | -12.96 | 1455.4 | 0 | ^a^NA | 0.99 |
| ^a^Estimate of 95% CI lower limit not able to be estimated due to small sample size. | | | | | |

| **eTable 7. SARS-CoV-2 Exposure and MCHAT-R Positive Screenings in Female and Male Subgroups (*COMBO-RSCH)*** | | | | | |
| --- | --- | --- | --- | --- | --- |
| **Predictor** | **B** | **SE** | **Odds Ratio** | **95% CI** | **P-value** |
| ***Females (n=160)*** |  |  |  |  |  |
| SARS-CoV-2 Infection in pregnancy | -0.34 | 0.70 | 0.64 | 0.15-2.38 | 0.52 |
| Age at M-CHAT administration (months) | -0.34 | 0.18 | 0.71 | 0.48-1.0 | 0.05 |
| Maternal age at delivery | -0.25 | 0.08 | 0.78 | 0.65-0.90 | 0.003 |
| Insurance (Medicaid) | 0.34 | 0.79 | 1.39 | 0.31-7.28 | 0.67 |
| Gestational Age | -0.22 | 0.15 | 0.80 | 0.59-1.08 | 0.13 |
| Maternal Race – American Indian or Alaska Native | -14.5 | 2796.04 | 0 | ^a^NA | 0.99 |
| Maternal Race – Asian | -14.47 | 1648.96 | 0 | ^a^NA | 0.99 |
| Maternal Race – Black or African American | -0.05 | 0.93 | 0.95 | 0.13-5.61 | 0.96 |
| Maternal Race – Declined | -1.14 | 1.37 | 0.03 | 0.01-3.42 | 0.41 |
| Maternal Race – White | -0.07 | 0.88 | 0.93 | 0.16-5.29 | 0.93 |
| Maternal Ethnicity - Declined | -0.45 | 1.42 | 0.64 | 0.02-9.71 | 0.75 |
| Maternal Ethnicity – Not Hispanic or Latino | -0.87 | 0.89 | 0.42 | 0.07-2.43 | 0.33 |
| Maternal Ethnicity – Unknown | -14.90 | 3956.18 | 3.36-07 | ^a^NA | 0.99 |
| ***Males (n=199)*** |  |  |  |  |  |
| SARS-CoV-2 Infection in pregnancy | 0.48 | 0.48 | 0.62 | 0.23 - 1.52 | 0.32 |
| Age at M-CHAT administration (months) | -0.21 | 0.29 | 0.81 | 0.42 - 1.28 | 0.46 |
| Maternal age at delivery | -0.05 | 0.04 | 0.95 | 0.87 -1.03 | 0.26 |
| Insurance (Medicaid) | 0.03 | 0.51 | 1.03 | 0.38 – 2.82 | 0.95 |
| Gestational Age | -0.36 | 0.14 | 0.70 | 0.52-0.92 | 0.01 |
| Maternal Race – American Indian or Alaska Native | 15.18 | 1455 | 0 | ^a^NA | 0.99 |
| Maternal Race – Asian | 1.71 | 0.97 | 5.54 | 0.78 – 37.69 | 0.08 |
| Maternal Race – Black or African American | 0.12 | 0.74 | 1.13 | 0.24-4.60 | 0.87 |
| Maternal Race – Declined | -0.41 | 0.72 | 0.66 | 0.15-2.60 | 0.57 |
| Maternal Race – White | 0 | 0.54 | 1.00 | 0.35-2.89 | 0.99 |
| Maternal Ethnicity - Declined | 0.27 | 0.69 | 1.30 | 0.35-4.99 | 0.70 |
| Maternal Ethnicity – Not Hispanic or Latino | -0.58 | 0.56 | 0.56 | 0.18-1.68 | 0.30 |
| ^a^Estimate of 95% CI lower limit not able to be estimated due to small sample size. | | | | | |
